# Minimizing school disruption under high incidence conditions due to the Omicron variant in early 2022

**DOI:** 10.1101/2022.02.04.22270473

**Authors:** Elisabetta Colosi, Giulia Bassignana, Alain Barrat, Bruno Lina, Philippe Vanhems, Julia Bielicki, Vittoria Colizza

## Abstract

As record cases due to the Omicron variant are currently registered in Europe, schools remain a vulnerable setting suffering large disruption. Extending previous modeling of SARS-CoV-2 transmission in schools in France, we estimate that at high incidence rates reactive screening protocols (as currently applied in France) require comparable test resources as weekly screening (as currently applied in some Swiss cantons), for considerably lower control. Our findings can be used to define incidence levels triggering school protocols and optimizing their cost-effectiveness.

---

Countries in Europe are currently suffering disruptions in schools due to the exceptionally high rates of Omicron incidence in the community and in particular in children^1^. School protocols are put under stress by requiring repeated quarantines or leading to large and sudden testing demand for children, overloading saturated surveillance systems^2,3^. Through modelling, here we compare school protocols in terms of resource peak demands, infection prevention, and reduction of schooldays lost, specifically under the high incidence conditions due to the Omicron variant.

## Modelling SARS-CoV transmission in schools and school protocols

We extended a stochastic agent-based model of SARS-CoV-2 transmission at school presented in detail by Colosi et al.^4^ (see Supplementary Information, SI). Infection progression includes prodromic, clinical and subclinical disease stages, informed from empirical distributions, and accounted for children’s lower susceptibility and transmissibility^5,6^.

Next to symptomatic testing, we simulated the reactive protocol currently applied in France, requesting an anterior nasal LFD test at days D0, D2, and D4 to the class of the detected case^7^. We compared it to two regular screening strategies, with weekly (as in the Baselland canton, Switzerland^8^) and semiweekly frequencies, assuming 75% (50-100%) adherence. Finally, we considered the reactive class quarantine after case detection, currently applied in Italian pre-primary schools^9^. Case isolation lasted 7 days.

The model was informed with time-varying and age-dependent test sensitivity, yielding estimated 67% peak sensitivity for asymptomatic children in nasal LFD tests and 96% in salivary PCR tests^10^ (SI). We also explored a lower peak sensitivity of 55%.

## Omicron epidemiological scenarios and test needs

We considered the circulation of the Omicron variant, assuming 20% protection from infection from prior variants^11^, an intrinsic transmissibility advantage of 30%-80% relative to Delta^12^, and a shorter incubation period^12,13^. The advantage is applied to the within-school transmissibility of previously circulating variants that we inferred in prior work from observed prevalence in French schools^4^ (SI).

Following current vaccine coverage in France, we considered that all teachers completed the primary vaccination (with estimated vaccine effectiveness against symptomatic disease from Omicron infection VE=50% at 3 months after the second dose^14^), with 50% of them having received the booster (VE=70% within the first 4 weeks^14^). In a first analysis, we assumed no vaccinated children (French coverage <3% by mid-January^15^).

The simulated Omicron wave captures the dynamics reported by community surveillance incidence in primary school students^15^ (Figure 1A). The reactive protocol marginally reduces the peak, whereas regular screening flattens more substantially the curve. The number of tests required by the reactive protocol increases along the wave, with a predicted peak demand of 0.45 (95% bootstrap CI 0.44-0.48) tests per student per week at an incidence close to 7,000 cases per 100,000 (Figure 1C). Test demand instead decreases in the regular protocols because fewer students are present in class after the peak of infections due to isolation (0.457 (0.457-0.461) tests in the weekly and 0.97 (0.97-0.98) in the semiweekly screening). Higher incidence conditions can lead to a larger demand of tests by the reactive protocol compared to the weekly screening (Figure 1BD).

**Figure 1.**
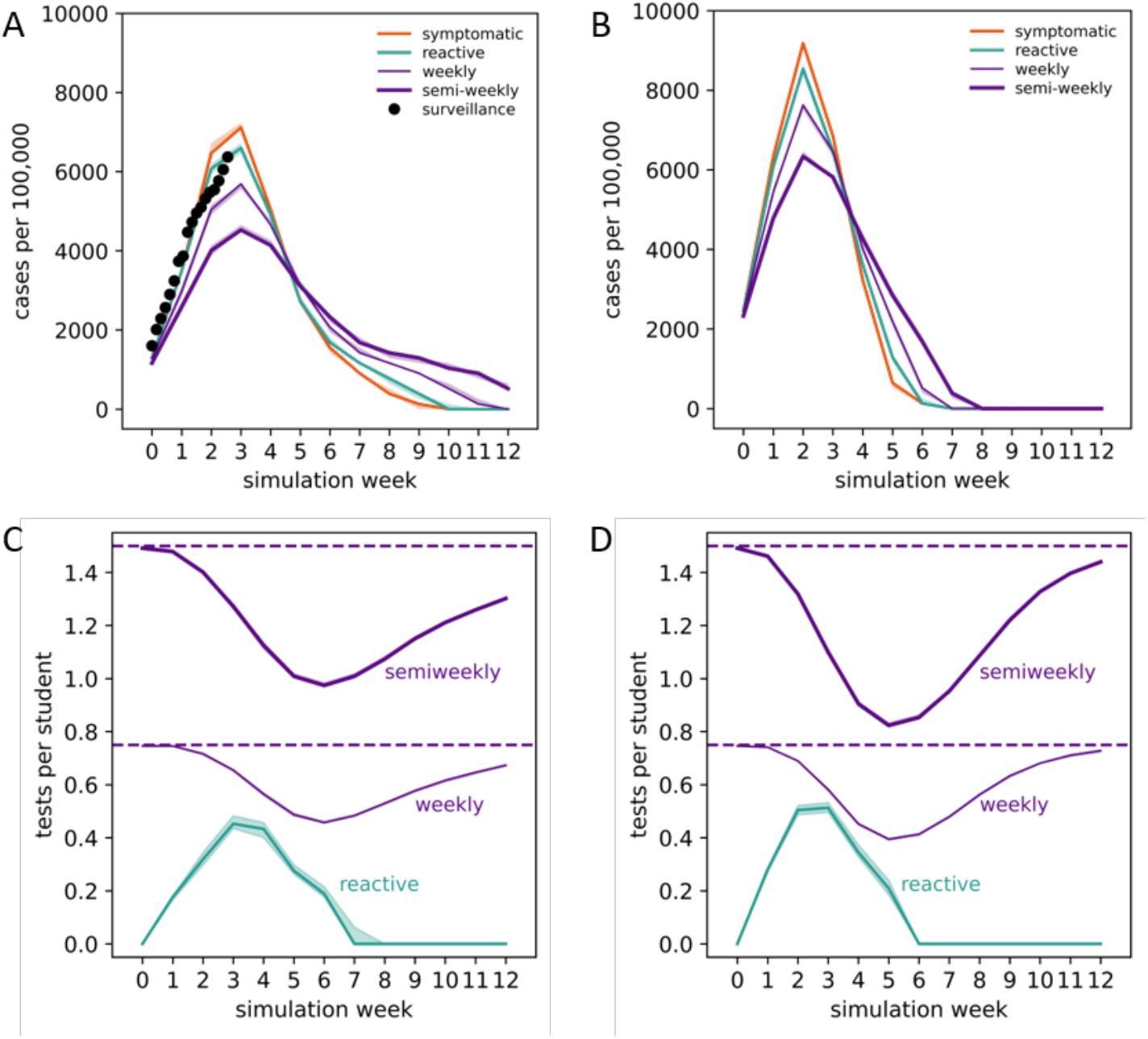
Incidence and number of tests per student over time under different school protocols. A,B: simulated weekly incidence expressed in number of cases in students per 100,000 over time for lower (A) and higher (B) introduction conditions, for different protocols. The left plot also shows the reported incidence in the 6-10y age class in France in the period 02-18/01/2022^15^. C,D: average number of tests per student over time for reactive and regular protocols under the epidemic conditions illustrated in the top panels. The horizontal dashed lines indicate the theoretical values of the demands in number of tests per student in the weekly and semiweekly screening (i.e. imposed by 75% adherence and by the frequency). Results are obtained considering the use of nasal LFD tests in both reactive and regular screenings. Shaded areas around the curves correspond to 95% bootstrap confidence intervals.

## Resource needs vs. impact on cases and schooldays lost

To evaluate how to best use resources, we extended the analysis of Figure 1 to a larger set of Omicron wave scenarios with varying peak incidence (SI). For increasing values of the incidence rate, from approximately 4,000 to 10,000 cases per 100,000, the number of reactive tests per student-week increases from 0.25 to 0.61 (Figure 2A). However, these tests would have marginal control of the viral circulation at school, reducing the peak of the wave by <10% (Figure 2B). For incidence up to 7,100 cases per 100,000 in absence of interventions, weekly screening would lower the peak by >20% while requiring <0.46 tests per student-week at peak (Figure 2AB). The same impact could be achieved even for higher incidence rates (<10,100 cases per 100,000) by doubling the frequency. Similar results are obtained considering the reduction of the epidemic size of the full wave (SI).

**Figure 2.**
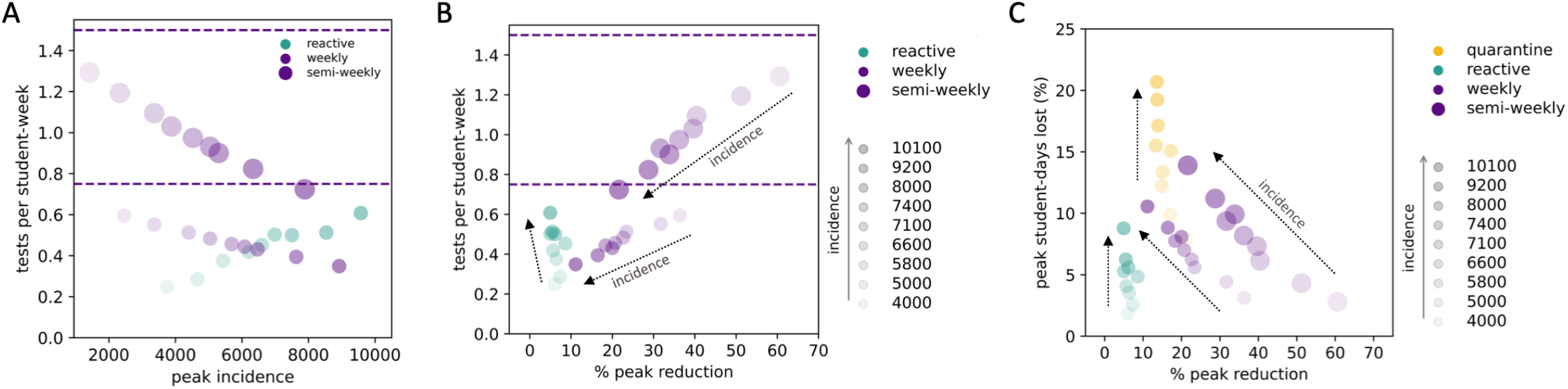
Test needs and schooldays lost vs. peak reduction at varying peak incidence rates. A: Demand in the number of tests per student-week at peak as a function of the peak incidence (cases in students per 100,000) for the reactive, weekly, and semiweekly protocols. The horizontal dashed lines indicate the theoretical values of the number of tests per student in the weekly and semiweekly screening (i.e. imposed by 75% adherence and the frequency). Dots reduce their transparency for increasing incidence. B: Demand in the number of tests per student-week at peak as a function of the percentage of peak reduction achieved by each protocol compared to symptomatic testing (i.e. in absence of interventions). The horizontal dashed lines are as in panel A. Dots transparency code is the same as in panel A. C: Peak percentage of student-days lost as a function of the percentage of peak reduction achieved by each protocol; both quantities are computed with respect to symptomatic testing. The reactive quarantine of the class is shown as an additional protocol. In panels B and C: incidence values in the legend refer to peak incidence of symptomatic testing (i.e. in absence of interventions); the corresponding values for each protocol are plotted in panel A; arrows are shown as guide to the eye.

Student-days lost remain below 11% with reactive and weekly screening, whereas reactively closing the class could lead to >20% of absence per student, compatible with observations^3^ (Figure 2C). Findings are robust against changes in booster coverage in teachers, in Omicron transmissibility and incubation period (SI). Higher detection rates would penalize the reactive screening, due to an increase in test demand while control remains limited (SI).

## Impact of test type, adherence, vaccination of children

Changing from nasal LFD tests to salivary PCR tests would improve the reactive strategy from 9% to 14% peak reduction if results are available after 12h (Figure 3A). Regular testing is instead mainly affected by adherence (Figure 3B). Vaccinating children would provide a collective benefit in reducing viral circulation at school. Assuming the vaccine effectiveness estimate for adults after 2 Pfizer doses (VE=60%)^14^, the peak would be reduced by 30%-45% for 40% to 60% coverage with the reactive screening, and by 35%-50% with weekly screening, compared to no vaccination (Figure 3C). For VE=20%, accounting for waning^14^, the reductions would be in the range 19-28% and 22-33% with the reactive and weekly screening, respectively.

**Figure 3.**
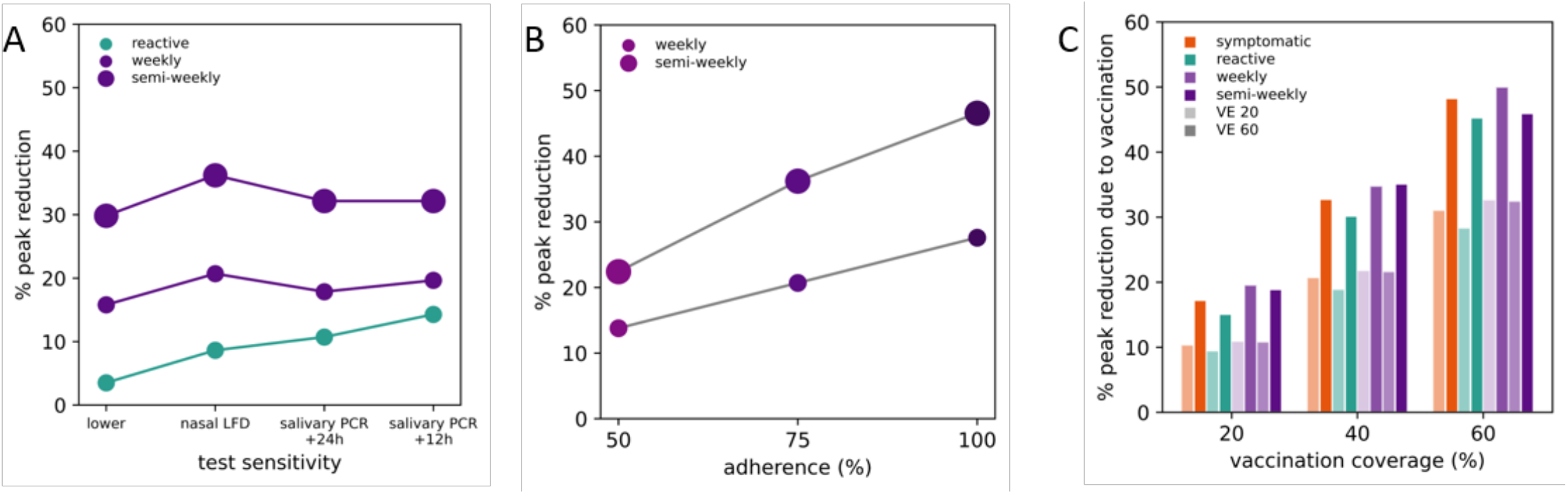
Impact of test sensitivity, adherence to regular screening, and vaccination. A: Percentage of peak reduction achieved by each protocol compared to symptomatic testing (i.e. in absence of interventions) as a function of the test sensitivity^10^ and of the delay in returning the results (+12h, +24h for PCR tests). The lower value corresponds to 55% peak sensitivity. B: Percentage of peak reduction as a function of adherence to regular screening. C: Reduction (%) in the peak incidence for each protocol due to vaccination in children, for different vaccination coverages. Vaccine effectiveness against symptomatic disease is set to VE=60% (solid bars), assuming the estimate of two Pfizer vaccine doses against Omicron obtained in adults^14^, and VE=20% (transparent bars), corresponding to the waned effectiveness estimated 3-4 months after the second dose^14^. Results of all panels refer to the Omicron wave shown in Figure 1A.

## Discussion

Given the high incidence rates currently recorded in Europe due to the Omicron variant^1^, our study shows that reactive screening strategies in schools, as employed in France, would require a similar number of tests per student per week compared to weekly screening, but achieve a lower epidemic control. The protocol requesting three tests in less than a week for case contacts in French primary schools led to large disruption in January 2022, in terms of logistics, resources, and impact on surveillance capacity^2^. We estimate that the same resources would be more efficiently used by weekly screening schools, reaching 20-40% peak reduction for incidence rates up to the values currently registered in France^15^, compared to the marginal reduction (<10%) of reactive screening.

Other countries opted for systematically screening schools against SARS-CoV-2 transmission, supported by numerical evidence^4,16–19^. Authorities in Baselland (Switzerland) offered weekly salivary PCR tests to all schools since March 2021. Prior to making participation mandatory in 2022, recorded adherence was on average >75%^8^. Proactively screening also has the advantage of planning resources in advance, contrary to reactive screening subject to sudden peak demands and potential shortages. This was reported to help simplifying the logistics of test-to-stay strategies in pilot weekly screenings implemented in a number of pre-primary and primary schools in the Auvergne-Rhône-Alpes region in France. Preliminary unpublished empirical estimates from these screenings also suggest a reduction of cases in December 2021 compared to the reactive strategy, in line with model predictions.

The now widespread use of nasal antigenic tests makes repeated self-testing possible without loss in efficiency, as lower sensitivity is compensated by promptness of results and high frequency. It would also limit the high rates of absence from school due to reactive class closures that are predicted by the model and currently reported^3^.

As European countries approach the peak of the Omicron wave, these findings can be used to tune the response by defining incidence levels triggering protocols adapting to the decreasing phase of the wave, according to the established objectives. Systematically screening schools remains the optimal test-to-stay strategy, reducing peak incidence rates in children, and thus their consequences on hospitalizations and long COVID^20^, while limiting school disruption and resources. Large vaccination coverage in children contributes to mitigate high viral circulation, making schools safer. Current coverage remains however low in children in several European countries (14% median coverage for first dose vaccination by week 4 2022; SI).

Our study has limitations. We did not fit the model to the Omicron wave in a specific country, as we aimed to evaluate protocols’ resources and impact in a range of high incidence conditions experienced across countries in Europe. Nonetheless, the model follows the observed dynamics in France. With remaining uncertainty on Omicron features, we assumed that its higher spreading rate was mainly due to immune evasion^12^, in line with observations from household studies^21^. Considering a higher intrinsic transmission led to similar conclusions. We did not consider immunity waning over time, but tested waned vaccine effectiveness.

A large demand in tests results from reactively screening schools in high incidence conditions. Comparable resources could be more efficiently used in a proactive screening strategy to mitigate the peak.

## Data Availability

All data produced in the present work are contained in the manuscript

## Acknowledgments

We thank Thomas Bénet for useful discussions. This study was partially funded by: ANR projects COSCREEN (ANR-21-CO16-0005) and DATAREDUX (ANR-19-CE46-0008-03); ANRS-MIE project EMERGEN (ANRS0151); EU H2020 grants MOOD (H2020-874850) and RECOVER (H2020-101003589); EU HORIZON grant VERDI (101045989); REACTing COVID-19 grant.

